# The Fault in Our Sets: A Mixed Methods Analysis of Clinical Value Set Errors

**DOI:** 10.1101/2025.02.27.25323054

**Authors:** Laura Zahn, Hasan Ahmad, Dean Sittig, Elise Russo, Briah Koh, Arianna Nimocks, Adam Wright

**Affiliations:** Department of Biomedical Informatics, Vanderbilt University Medical Center, Nashville, TN, USA; Parkview Health, Fort Wayne, IN, USA; Department of Clinical and Health Informatics, University of Texas Health Science Center, Houston, TX, USA; Marnix E. Heersink School of Medicine, University of Alabama, Birmingham, AL, USA; Bloomberg School of Public Health, Johns Hopkins University, Baltimore, MD, USA; Department of Medicine, Vanderbilt University Medical Center, Nashville, TN, USA

**Author notes:** **Corresponding Author:** Adam Wright, PhD, Department of Biomedical Informatics, Vanderbilt University Medical Center, 2525 West End, Nashville, TN, USA.

## Abstract

**Objective:** To characterize clinical value set issues and identify common patterns of errors.

**Materials and Methods:** We conducted semi-structured interviews with 26 value set experts and performed root cause analyses of errors identified in electronic health records (EHRs). We also analyzed a random sample of user-reported issues from the Value Set Authority Center (VSAC), developing a categorization scheme for value set errors. Additionally, we audited medication value sets from three sources and assessed the impact of value set variations on a clinical quality measure within Vanderbilt’s Epic system.

**Results:** Interviews highlighted ongoing difficulties in value set identification, creation, and maintenance, with significant consequences for clinical decision support (CDS), quality measurement, and patient care. Content analysis indicated that 42% of errors involved missing codes, 14% included extraneous codes, and 40% arose from misinterpretations of value set intent; 72% of these errors were present at creation. The audit revealed errors in 50% of medication value sets, predominantly omissions. The impact analysis demonstrated that value set selection altered a clinical quality measure’s outcome by 3- to 30-fold.

**Discussion:** Value set errors are widespread and arise from a delineable set of causes. Characterizing patterns of errors allowed us to identify best practices and potential solutions to minimize their frequency.

**Conclusion:** Better tools for finding, authoring, auditing and monitoring value sets are urgently needed.

## INTRODUCTION

Longitudinal electronic health records (EHRs) contain a range of clinical data, including medications, laboratory test results, diagnoses, orders, and clinical observations.^1^ Much of this data is structured and coded using standard terminologies such as Logical Observation Identifiers, Names, and Codes (LOINC), Systematized Nomenclature of Medicine – Clinical Terms (SNOMED CT), or RxNorm (a standardized nomenclature for clinical drugs).^2^ There are many benefits of coded data, including semantic interoperability, clinical decision support, and clinical quality measurement.^3^ In many cases, users of clinical data may need to refer to groups of related concepts. To support this need, informaticians create value sets, which the National Library of Medicine (NLM) defines as “lists of codes and corresponding terms that define clinical concepts.”^4^ For example, a value set for “Diabetes Mellitus, poorly controlled,” could consist of a list of SNOMED and ICD-10 diagnosis codes that represent poorly controlled diabetes (e.g., SNOMED CT concept 443694000 Type II diabetes mellitus uncontrolled or ICD 10 code E11.11 Type 2 diabetes mellitus with ketoacidosis with coma), while a value set for “lipid-lowering agents,” could include RxNorm codes for drugs designed to reduce a patient’s lipid levels (e.g. RxCUI 617310 atorvastatin 20 MG Oral Tablet).^4^ Value sets are commonly used in clinical decision support (CDS). For example, an alert that “suggests *pneumococcal vaccination* in patients over 65, or with *high-risk medical conditions*” uses two value sets (i.e., pneumococcal vaccination variants and high-risk medical conditions), while a clinical quality measurement, such as, “what proportion of patients with *hyperlipidemia* have a recent *LDL*?” also uses two sets (i.e., one for the concept hyperlipidemia, and the other for a variety of LDL test identifiers). Value sets also support many other use cases including reporting,^5,6^ interoperability,^5,6^ registries,^7^ research,^8,9^ data display,^10^ and summarization.^11^

When a value set is needed for a particular application, individuals can either select an existing value set or create their own. The Value Set Authority Center (VSAC), provided by the National Library of Medicine (NLM) in collaboration with the Office of the National Coordinator for Health Information Technology (ONC) and Centers for Medicare and Medicaid Services (CMS), is a repository of 15,214 current public value sets (as of July 2024) and is intended to be an authoritative resource. However, VSAC does not, itself, review, verify, or audit value sets, and instead leaves this to its 97 stewards, including the Joint Commission, CMS, Mathematica, and various medical specialty societies.^12^ Healthcare organizations also develop and maintain value sets locally, using a variety of processes and home-made tools (often as simple as a spreadsheet).^13–18^

Controlled medical vocabulary management is inherently complex,^3,19–22^ and creating accurate, up-to-date value sets is particularly challenging. Value set authors must take vague concepts (“at risk for pneumococcal infection”) and render them as precise value sets where any single code is either in or out of the value set.^23–27^ These authors may be expert terminologists, CDS or clinical quality measure (CQM) authors, clinical subject matter experts (who may or may not have informatics training), or software developers (with no clinical or informatics training). These authors must understand the needs of the targeted value set user and anticipate the needs of other potential value set users, as well as the behaviors of the healthcare providers that are entering the codes (like diagnoses or problem list entries) that will be compared to those in the value set.^28^ Moreover, clinical evidence and practice shift over time^29^ as new drugs are introduced or new indications are found for existing drugs;^30^ new diseases are identified or their names change;^31,32^ or clinical procedures are added, changed, or reorganized.^33–37^

Value set sharing and reuse has long been promoted in the field, despite demonstrated problems of bias and inaccuracy in value sets shared on public repositories.^6,38–40^ For example, an analysis of 32 VSAC value sets for 12 chronic conditions found substantial discrepancies between value sets, including a total of 189 potentially problematic codes frequently used in EHRs.^41^ Value set errors can negatively impact the quality of patient care. In our previous work, we found value set errors to be the most common cause of CDS malfunctions.^15^ CDS malfunctions put patients at risk and have led to patient harm and death.^15,16^ Even when CDS malfunctions are intercepted by astute clinicians, they contribute to alert fatigue and undermine clinician trust in CDS and quality measurement.^42^ Value set errors can also cause inaccuracies in CQMs.^43^ In this paper, we present a case series of value set errors that occurred in practice at academic hospitals and then report the results of a content analysis of randomly selected user-submitted value set issues. We conclude with a series of recommendations for improving value set creation and maintenance processes.

## METHODS

### Subject Matter Expert Interviews

We conducted 21 semi-structured interviews with 26 value set experts from six academic healthcare systems, as well as First Databank, MD Partners, and members of the electronic clinical quality measure (eCQM) Value Set Workgroup (including personnel from Mathematica). Interview topics included value set creation processes, identification challenges, usage patterns, and error examples from personal experience. Thematic analysis identified common challenges and error patterns.

### Case Series

We compiled a case series of value set errors from academic medical centers, identified via user reports, CDS issue analyses, and experts interviews. Each case was reviewed to determine error origins and clinical consequences.

### Issue Review and Content Analysis

The Assistant Secretary for Technology Policy*/*Office of National Coordinator for Health Information Technology (ASTP/ONC) and the Centers for Medicare and Medicaid Services (CMS) maintain a Jira-based, eCQM issue tracking system, where value set users can report issues found in VSAC and used in quality measurement programs.^44^ Since eCQMs rely on value sets to define data elements, not all the issues reported on Jira are related to value sets. For example, users may report issues with timing of data elements, test scripts, or even their account or password. We reviewed a random sample of issues reported on Jira between January 1, 2019 and December 31, 2020, and evaluated over 1000 until we identified 100 issues specifically related to value sets for further study. The research team categorized errors by type (missing codes, extra codes, both, or neither) and timing (congenital-present at creation or acquired-post-creation), using ticket descriptions and value set histories. Two authors (L.Z. and H.A.) inductively derived error sources from Jira tickets, finalizing categories through consensus with the entire study team.

### Clinical Audit

We conducted a clinical audit of medication value sets from three sources: 1) VSAC’s 2025 eCQMs (all 86 available), 2) CQM value sets from VUMC (a random sample of 100 selected, sourced from measure programs) and 3) CDS-related value sets in at VUMC (a random sample of 100 selected, all locally developed). Each value set was reviewed in context for missing medications or erroneously included medications.

### Impact Analysis

For the impact analysis, we selected an eCQM and its associated CDS involving prescription of beta blockers after hospital discharge for secondary prevention of myocardial infarction (MI), we identified all potentially related value sets in Vanderbilt’s Epic installation. We then evaluated the number of patients with the diagnosis codes in each diagnosis value set (the “denominator” population of eligible patients), and evaluated the number and percentage of patients who were taking a beta blocker for each pair of diagnosis (denominator) and medication (numerator) value sets.

The Vanderbilt University Medical Center (VUMC) Institutional Review Board gave ethical approval for this work.

## RESULTS

### Subject Matter Expert Interviews

Experts highlighted difficulties in selecting a value set to reuse due to overlapping names and unverifiable accuracy, often prompting redundant local creations. They noted inadequate tools for maintaining value sets and detecting errors, with false positives (extra codes) easier to spot via user feedback than false negatives (missing codes). A need for advanced comparison and monitoring tools was widely endorsed.

### Case Series

**Case 1.** Carvedilol is a beta-blocker which also has alpha-blocker activity. Some drug databases do not include it in their beta-blocker drug class but, instead, in a separate class of “mixed alpha- and beta-blockers.” Many organizations have built CDS alerts which suggest prescribing a beta-blocker for patients who have an indication for one but are not currently prescribed one. In separate cases at different organizations, an inaccurate alert suggested prescribing a beta-blocker for a patient already taking carvedilol, and in at least one case, a patient received carvedilol in addition to another beta-blocker and experienced symptomatic bradycardia which required hospitalization. We described this issue in more detail in a prior paper.^13^

**Case 2**. A commercially developed postpartum hemorrhage (PPH) risk prediction model was used to drive CDS for PPH prevention and treatment. The model used a value set of SNOMED codes. SNOMED retired a “threatened abortion” code and replaced it with “threatened miscarriage”. The PPH value set was not updated to replace the retired code with the new code and became empty. As a result, the PPH model systematically underestimated the risk of PPH for women worldwide, failing to flag women at high risk of PPH. The error persisted for four years before being noticed by an astute analyst at a customer site.

**Case 3.** A teen girl sought pregnancy and sexually transmitted disease testing from her pediatrician. The patient requested that the results (or even the fact that she was being tested) not be shared with her parents, and the pediatrician assured her that they would be suppressed from the patient portal in accordance with state law. The EHR’s result release module used a value set to identify sensitive tests to suppress. Urine and blood pregnancy tests performed in the lab were included in the value set, but point-of-care office-based pregnancy tests were accidentally omitted. Her parents saw the negative pregnancy test result through the shared patient portal, deduced she was sexually active, and confronted her about being sexually active. The patient attempted suicide and was admitted to the hospital.

**Case 4.** The Joint Commission (TJC) and the Centers for Medicare and Medicaid Services (CMS) promulgated an electronic clinical quality measure (CQM): the proportion of patients with stroke discharged from the hospital on lipid-lowering therapy (CMS105v3/STK-6). The associated “Lipid-Lowering Agent RxNorm-based Value Set” (2.16.840.1.113883.3.117.1.7.1.217) released in VSAC contained the expected drugs, such as statins, ezetimibe and fenofibrate, but also incorrectly contained clopidogrel, an anti-platelet drug which does not reduce lipids. For 13 months, organizations that used the CQM incorrectly counted stroke patients discharged on clopidogrel as compliant with the quality measure. Some organizations also used the same value sets to drive CDS and reports, meaning that stroke patients needing lipid-lowering therapy may not have been identified and treated.

**Case 5.** A hospital built an alert for immunocompromised patients who had orders for live-virus vaccines, which are relatively contraindicated in this population due to the possibility of iatrogenic infection. The knowledge engineer who developed the diagnosis value set for “immunocompromise” included only codes for primary immunodeficiency (relatively rare) but not more common forms of acquired immunocompromise (like HIV, chemotherapy, or steroid use). Even the literal diagnosis “immunocompromise” was not included in the value set, putting patients at risk for receiving relatively contraindicated live-virus vaccinations.

**Case 6.** A set of CDS alerts fire when potassium chloride is ordered for patients with high potassium levels. The EHR had four codes for potassium results (whole blood vs. serum potassium levels, and lab-performed vs. point-of-care tests). The alert looked only at one of the code combinations, so it sometimes did not fire if patients had high potassium levels measured via a different type of potassium test and could also fire inappropriately if a previous value for the included potassium code was high, but there was a more recent normal potassium level associated with one of the code combinations missing from the value set.

**Case 7.** A value set for structural heart disease (conditions that affect the heart’s valves, walls, chambers, or muscles) contained unexpected diagnoses like “land mine injury” or “food refusal.” The value set had been exported from an EHR, edited, and reimported into a different environment of the same EHR. The two EHR environments had differences in internal ID numbers for some diagnosis codes, causing some of the imported entries to point to incorrect diagnosis codes. This caused an alert to fire inappropriately for certain patients with structural heart disease, who should have been excluded.

**Case 8.** After a case of patient harm, an alert was developed that reminds hospital-based physicians that a patient has an insulin pump so it can be used or turned off while the patient is in the hospital. A value set of insulin pump identifiers was created for use in the alert logic. As new insulin pumps were released, the value set was not updated, so the alert failed to fire for about 25% of patients with insulin pumps, putting them at risk for hypoglycemia, as physicians had started to depend on the alert.

### VSAC Issue Review

Our review of 100 value set issues reported in ONC’s Jira revealed the largest proportion of problems involved missing codes (42%) (Table 1). Forty percent of issues reviewed involved neither missing nor extra codes, meaning the value set contents were correct, but issues still arose, most often due to misunderstanding of the value set intent or purpose. A smaller proportion of reported issues involved extra codes, meaning contents included something that did not belong. Only four of the 100 issues reviewed involved both missing and extra elements. Most of the issues reviewed were determined to be congenital (72%), meaning the errors occurred at the point of value set creation, while the remaining (28%) were acquired over time as circumstances changed.

**Table 1.** Count of error types identified when reviewing a random sample of 100 ONC Jira reported issues.

| Type of error | Example | Count (n=100) |
| --- | --- | --- |
| Missing codes | A new flu shot was released, but the code was not added to the value set in time | 42 |
| Neither missing nor extra codes | User requests lower prophylactic dose forms be added to a value set for therapeutic doses of heparin. Request declined since the dose is not appropriate for the intent of the CQM measure (Anticoagulation Therapy for Atrial Fibrillation/Flutter). | 40 |
| Extra codes | Certain value sets had duplicate rows (e.g. contained the gender "female" twice) which caused some systems to not be able to read them if they had a uniqueness constraint | 14 |
| Both missing and extra codes | Value set for cancer diagnosis included three missing ICD-10 codes and five erroneous ICD-10 codes that needed removal | 4 |

Underlying sources of errors were inductively derived from Jira tickets and the resulting categories and overarching domains are shown in Table 2. The most common source of value set issues reported to ONC’s Jira was user confusion related to value set intent or contents (26%). Other common sources included the value set inappropriately using one coding system or aspect of the coding system (17%), and a simple oversight on the creator’s part leading to important exclusions (13%). Guideline changes were determined to be the cause for 11% of the issues reported, mostly due to value sets not being updated after the change and fewer resulting from users being unaware of a guideline change.

**Table 2.**
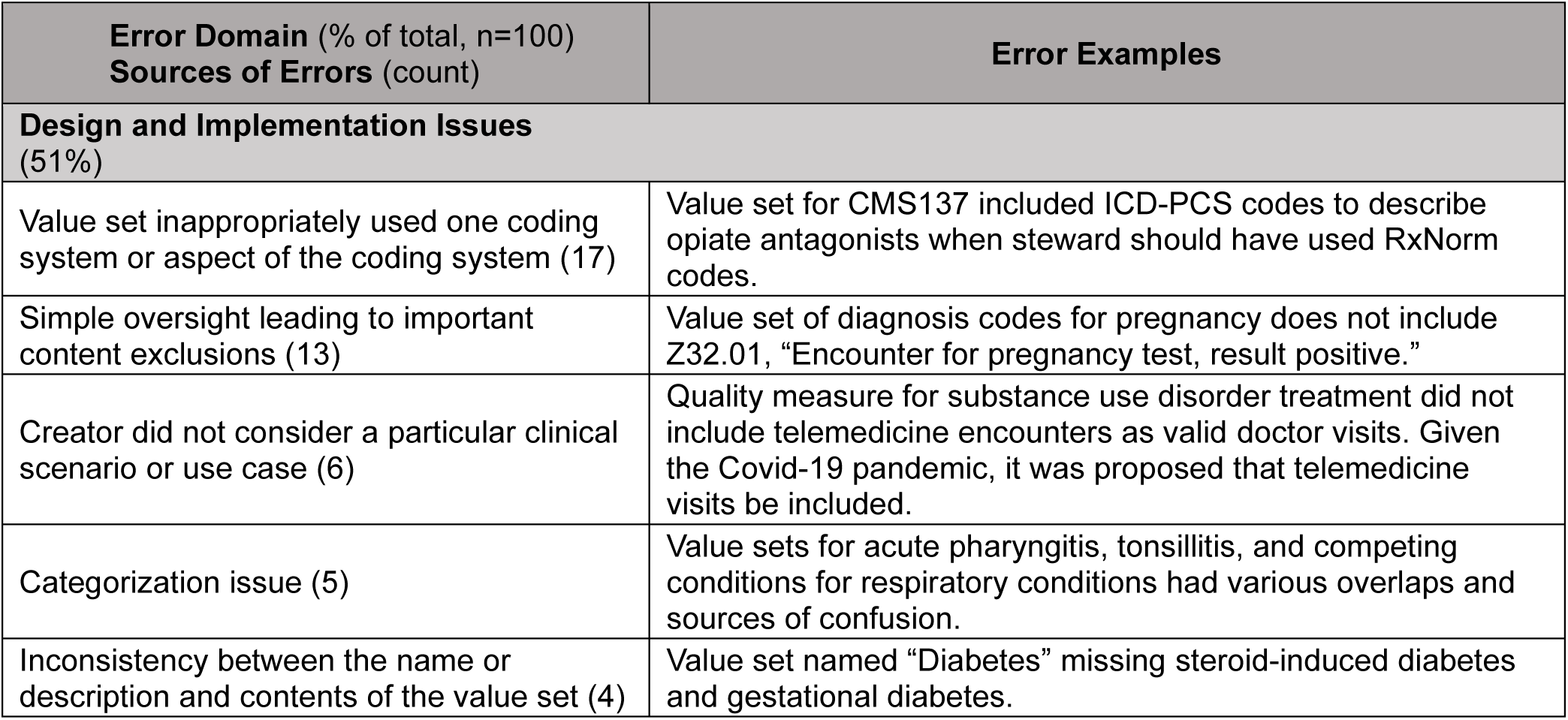
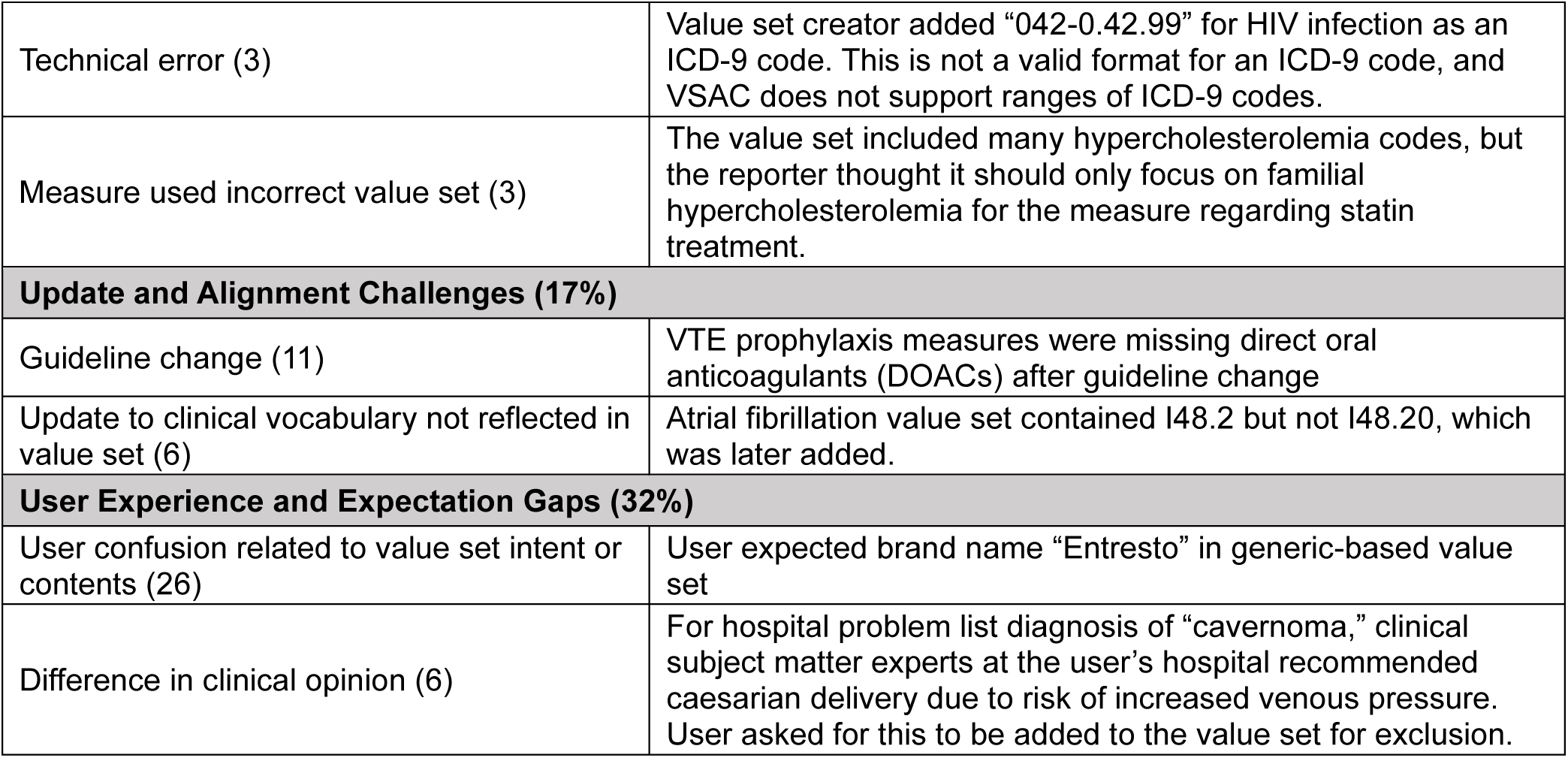
Sources of value set errors, definitions, and their count of Total Errors.

### Clinical Audit

Our clinical audit of medication value sets from three sources revealed errors were present in 50% of all value sets reviewed (Table 3). Missing items was the most frequent error type, though many value sets had both missing and erroneous inclusions.

**Table 3.** Audit of medication value sets from VSAC and VUMC used for clinical quality measurement or clinical decision support.

|  | **VSAC 2025 eCQM value sets** | **VUMC CQM Value Sets** | **VUMC CDS Value Sets** |
| --- | --- | --- | --- |
| **Creator** | CMS and VSAC stewards including AHA, Mathematica, TJC, NCQA | Sourced from VSAC, TJC and NCQA | Locally-developed at VUMC |
| **Authoritative** | Yes | Yes | No |
| **Usage** | CQMs for CMS’s inpatient and outpatient hospital reporting programs | Various CQM programs, including HEDIS, PQRS, TJC Core Measures | CDS tools, including alerts, scoring systems and predictive models |
| **Number reviewed** | 86 (all available) | 100 (random sample) | 100 (random sample) |
| **% with erroneous inclusion** | 21% | 18% | 24% |
| **% with missing item** | 41% | 57% | 43% |
| **Examples** | dextromethorphan (an antitussive) in a value set of antihistamines<br>metolazone missing from a diuretic value set<br>budesonide | dalteparin, an anticoagulant, in an antidepressant value set<br>doxycycline eyelid wash in a value set for streptococcal pharyngitis treatment | clozapine, the antipsychotic of choice for patients with Parkinson’s disease, in a value set for medications contraindicated in Parkinson’s |
|  | missing from a glucocorticoid value set | • cefazolin missing from a beta-lactam antibiotic value set<br>• newer agents such as PCSK9 inhibitors and bempedoic acid missing from a lipid-lowering agent value set | • ketoconazole missing from a warfarin DDI value set<br>• SGLT2 and DPP4 inhibitors missing from an antidiabetic medication value set |

### Impact Analysis

For a CQM and associated CDS involving prescription of beta blockers after hospital discharge for secondary prevention of myocardial infarction, our organization’s Epic installation includes 20 myocardial infarction value sets (the “denominator” population of eligible patients), with contents ranging from 39 to 416 diagnoses codes, and 10 beta blocker medication value sets, with contents ranging from 67 to 386 unique medications. Similar to our other findings, many of the associated value sets were missing important contents while others included inappropriate codes (Figure 1). To determine the percentage of patients having a MI and taking a beta blocker for the CQM, there were 200 possible value set combinations, resulting in a wide range of eligible patients: minimum 5,559 patients (25.7%) and maximum 10,806 patients (77.3%) (Figure 2).

**Figure 1.**
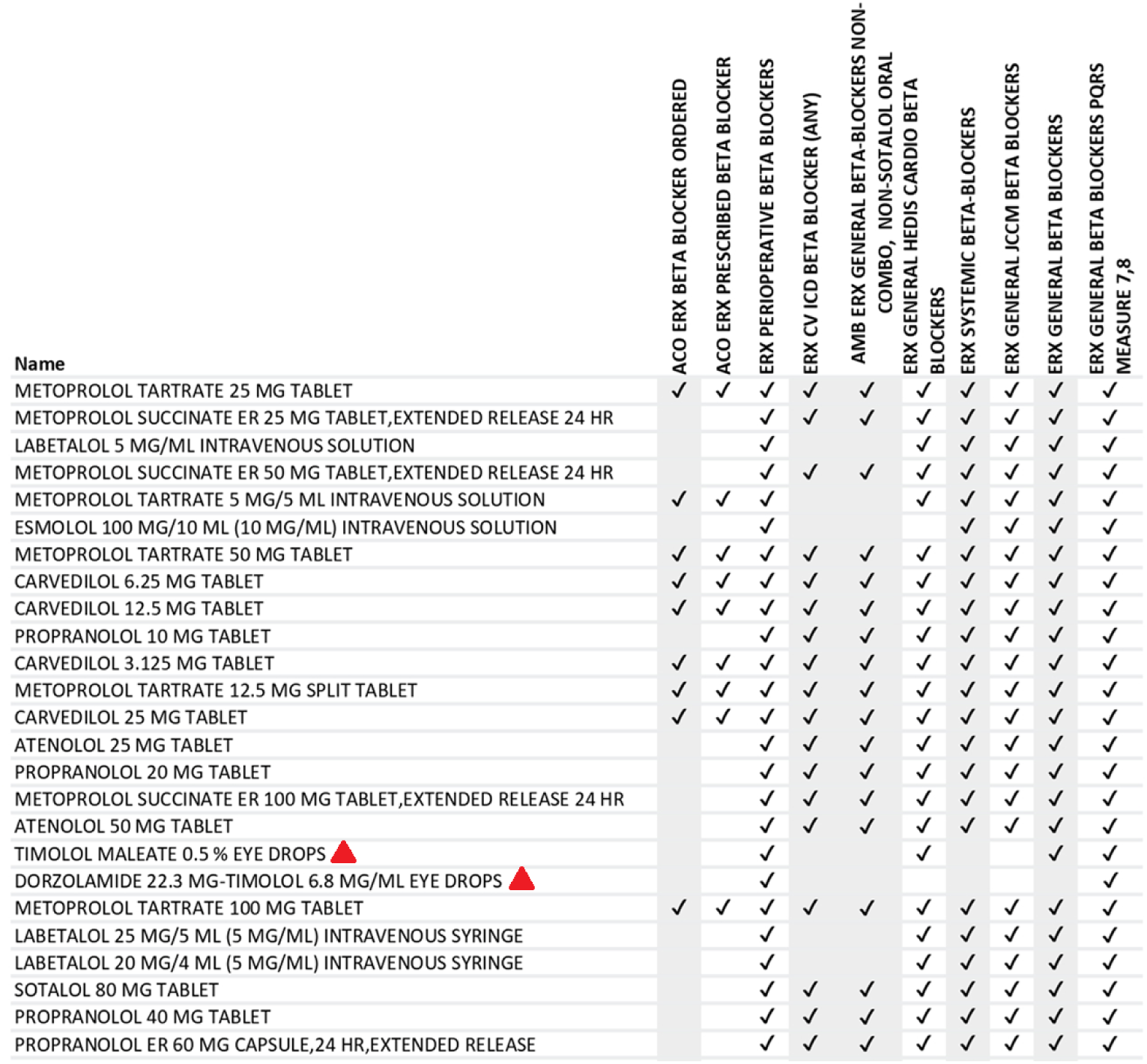
Table showing which of the top 25 beta blocker medications prescribed are included (indicated with check marks) in the 10 different beta blocker value sets. Some value sets were missing important contents while others included eye drop formulations not relevant to this context.

**Figure 2.**
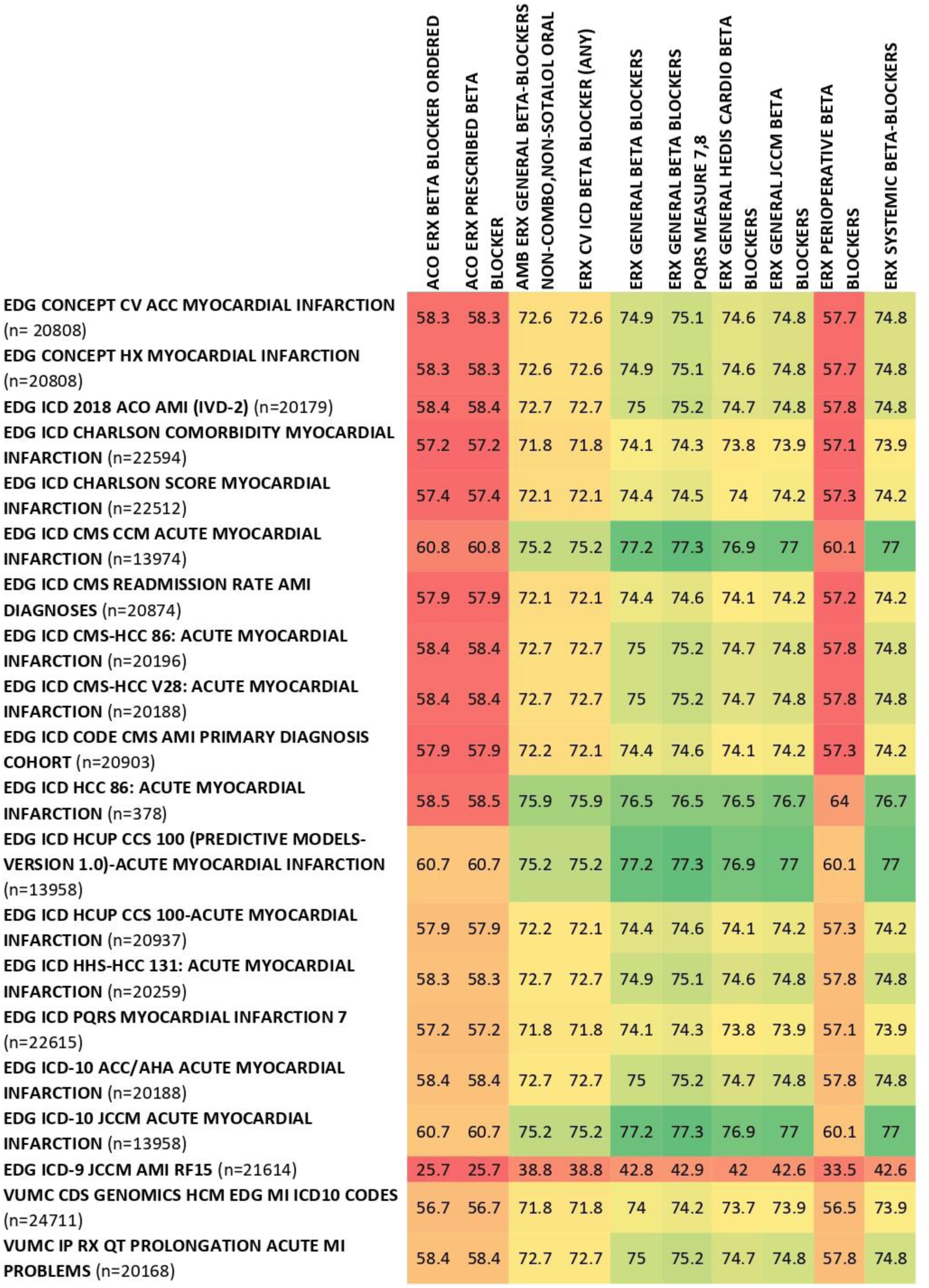
Heat map showing the percentage of patients having MI and taking a beta blocker using various combinations of value sets.

## DISCUSSION

Our research reveals that value set errors are not isolated incidents but systematic problems with far-reaching consequences. The case series demonstrates how these seemingly technical errors directly impact patient care—from inappropriate medication prescriptions to confidentiality breaches leading to attempted suicide. Our clinical audit confirms this is not an isolated problem: half of all medication value sets contained errors, regardless of whether they were created by national authorities or local teams.

The magnitude of these errors is particularly alarming. Our impact analysis revealed that simply changing which value sets are used can alter clinical quality measurement results by 3- to 30-fold for the same patient population. In practical terms, this means a hospital could appear to have excellent performance or poor performance on the same quality measure depending solely on which value set was selected. These discrepancies affect not only quality reporting and financial reimbursement but also direct patient care—determining which patients receive interventions like beta blockers and which do not.

Our mixed methods study also characterized common patterns of value set creation, errors, and their sources. These results align with prior work, notably Williams et al. which identified similar creation and maintenance challenges.^3^ However, we extend these findings by quantifying error types and detailing clinical impacts, offering a nuanced taxonomy across three domains. While the impact of these errors can range widely in terms of severity, they offer useful lessons for improvement. By recognizing the patterns within these occurrences, we can develop better tools, systems, or solutions to minimize their frequency.

One pattern that became clear from our study is that most errors appear to be congenital, occurring at the time of value set creation. This highlights the difficulty in creating an accurate and complete value set from the start. Key challenges with value set creation include:

- Clinical medicine is complex, and accurately characterizing complex and varied human physiology and pathophysiology using a finite list of codes is inherently difficult.
- Clinical terminologies differ in granularity and scope.
- Users may code data differently than value set creators expect or may even code data incorrectly. In addition, value set creators may work in specialty societies, government, or third-party organizations, so they may choose obscure codes for value sets, that while technically correct, are not routinely used by clinicians to encode patient data in the EHR.
- Different healthcare organizations and providers may interpret and use codes differently.
- Current tools for developing and maintaining value sets are rudimentary – in many cases, simple spreadsheets are used.
- Creating value sets requires a mix of clinical expertise and informatics skills such as EHR configuration expertise, knowledge of terminologies, Boolean logic, concept modeling, and clinical knowledge management. It is uncommon for a single person to have all these skills.
- Value set creators are human and may make typographical errors, commit simple oversights, or fail to consider all possible scenarios or use cases.

A smaller but not insignificant number of errors evaluated in our study were acquired, indicating they arose over time as conditions changed. This is especially true for errors relating to “guideline changes” and “updates to clinical vocabulary” as it requires concerted effort to adequately maintain value sets after their creation. In addition, a clinical scenario may not be considered at the time of value set creation, but then becomes obvious after circumstances change, like the COVID-19 pandemic and the sudden and widespread adoption of telemedicine. Regular auditing and updating of existing value sets can help detect or prevent these issues, but, given the large number of value sets and terms in use, automated tools and processes for monitoring and maintaining value sets are needed.

Another pattern that became clear from our study is that many value set issues arise from misunderstandings of value set intent between creators and users of value sets. Oftentimes, it is not easy to understand the intended purpose of a value set from its name alone and “user confusion related to value set intent or contents” was the most common error source in our issue review. When a user aims to reuse an existing value set created by someone else, the contents must be analyzed closely to ensure they also fit the current user’s need. Misusing a value set can lead to critical exclusions and inappropriate inclusions with potentially harmful results. Better metadata, descriptions of the rationale for pertinent inclusions or exclusions, and tools for finding, visualizing, and comparing value sets are needed.

Through our research, including expert interviews and our review of issues reported, we identified some best practices to improve value set creation, maintenance, and user experience, which are summarized in Table 4 below.

**Table 4:**
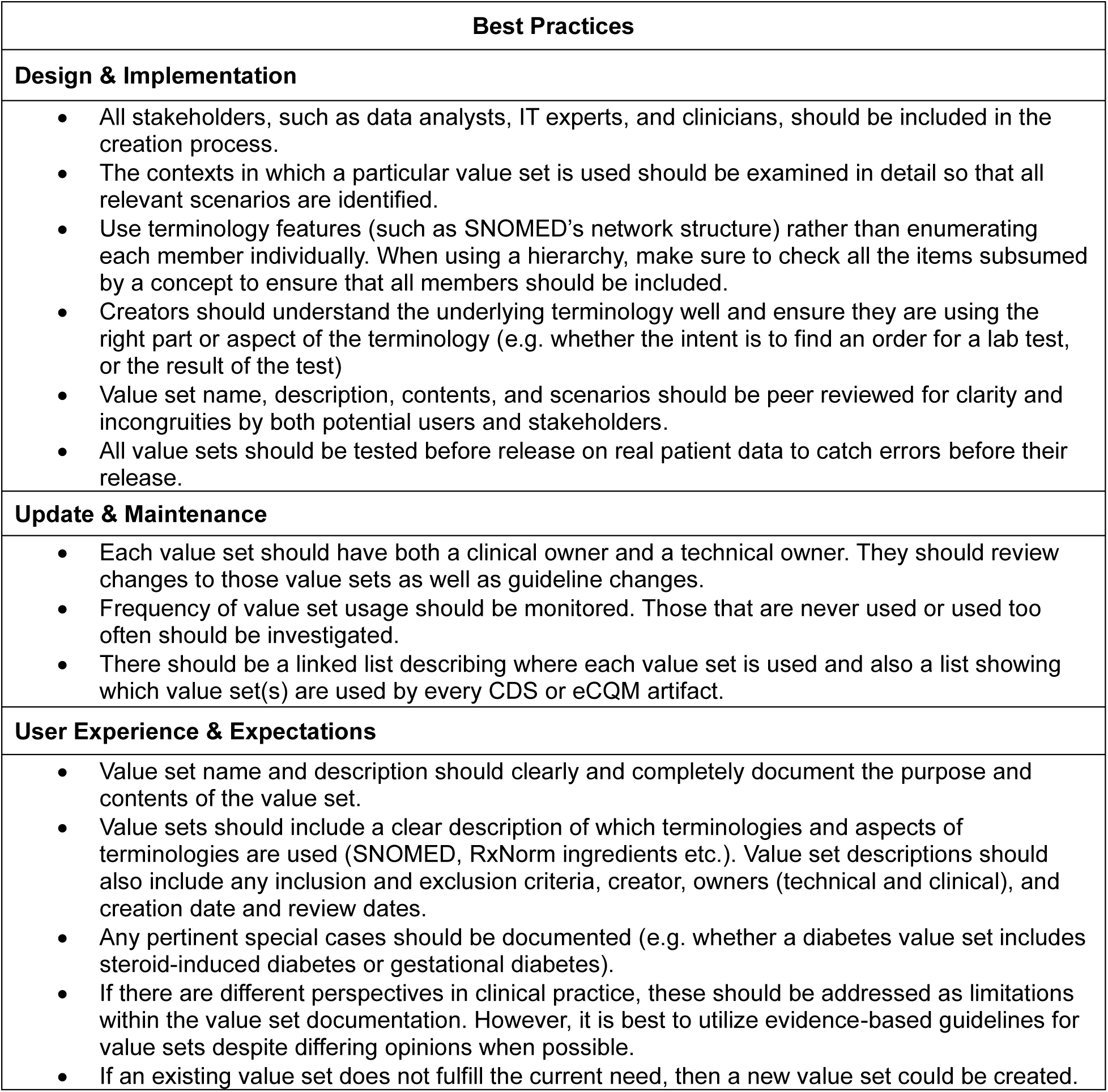
Best Practices.

### Limitations

Our study has several limitations. First, our case series was based on a convenience sample of cases, so future studies could use survey methods to gather a more representative sample. Second, although VSAC is the authoritative source of value sets, other organizations, including EHR vendors, healthcare organizations, and clinical content vendors also maintain value sets. It would be informative to study issues in these alternative value set sources. Finally, many value set errors have yet to be found. Since we focused on known issues, there may be subtler types of value set errors not detected as readily that would have been excluded in our sample.

### Future Work

Our work has highlighted the need for better tools to enable users to both find and compare the contents of different value sets. This should include the ability to sort value set contents by frequency of use based on real patient data. These better tools should also allow organizations to keep track of how different value sets are being used, such as for clinical decision support rules and clinical quality measures. When describing technical errors, we found that sometimes an invalid value set was uploaded to VSAC. Creation of a value set syntax validator could prevent this from occurring in the future.

We have also discussed the importance of regular maintenance and updates to value sets. Guideline developers should release value sets along with their guidelines and provide updates for both simultaneously to reduce value set errors. Expanded use of machine learning and large language models could lend well to supporting the process of value set creation and maintenance, while also catching any content exclusions, decreasing maintenance time, and ensuring updates occur in a more timely fashion.^45,46^ Artificial intelligence and large language models could help suggest updates to value sets rather than waiting for end users to report value set issues. For example, if there is a change, a maintenance tool could generate an alert that certain value sets need to be reviewed.

## CONCLUSION

Value set errors represent a significant patient safety issue that demands immediate attention. The current manual processes for creating and maintaining these critical clinical definitions are demonstrably inadequate, with errors present in half of all medication value sets we examined. Healthcare organizations must implement the best practices identified in this study to address each error category: Design and Implementation Issues, Update and Alignment Challenges, and User Experience and Expectation Gaps.

Informatics researchers should develop automated validation tools that can detect missing or inappropriate codes before they affect patient care. Healthcare systems should establish clear ownership and regular review processes for value sets. Finally, value set creators and stewards must improve documentation of intent and scope to prevent misunderstandings. Without these changes, the healthcare system will continue to experience preventable clinical errors, inaccurate quality measurements, and potential patient harm from what appears to be a purely technical issue but has profound clinical implications.

## Data Availability

All data produced in the present study are available upon reasonable request to the authors.

## REFERENCES

1. Vera Ehrenstein, Hadi Kharrazi, Harold Lehmann, Casey Overby Taylor. Obtaining Data From Electronic Health Records. Tools and Technologies for Registry Interoperability, Registries for Evaluating Patient Outcomes: A User’s Guide, 3rd Edition, Addendum 2 [Internet] [Internet]. Agency for Healthcare Research and Quality (US); 2019 [cited 2024 Jul 1]. Available from: https://www.ncbi.nlm.nih.gov/books/NBK551878/

2. Oliver Bodenreider, Ronald Cornet, Daniel J. Vreeman. Recent Developments in Clinical Terminologies — SNOMED CT, LOINC, and RxNorm. Yearbook of Medical Informatics. 2018 Aug;27(1):129–139. PMCID: PMC6115234

3. Richard Williams, Evangelos Kontopantelis, Iain Buchan, Niels Peek. Clinical code set engineering for reusing EHR data for research: A review. Journal of Biomedical Informatics. 2017 Jun;70:1–13. PMID: 28442434

4. Code Systems and Tools [Internet]. U.S. National Library of Medicine; [cited 2024 Jun 10]. Available from: https://www.nlm.nih.gov/vsac/support/authorguidelines/code-systems.html

5. Deborah J. Cohen, David A. Dorr, Kyle Knierim, C. Annette DuBard, Jennifer R. Hemler, Jennifer D. Hall, Miguel Marino, Leif I. Solberg, K. John McConnell, Len M. Nichols, Donald E. Nease, Samuel T. Edwards, Winfred Y. Wu, Hang Pham-Singer, Abel N. Kho, Robert L. Phillips, Luke V. Rasmussen, F. Daniel Duffy, Bijal A. Balasubramanian. Primary Care Practices’ Abilities And Challenges In Using Electronic Health Record Data For Quality Improvement. Health Affairs (Project Hope). 2018 Apr;37(4):635–643. PMCID: PMC5901976

6. Sigfried Gold, Andrea Batch, Robert McClure, Guoqian Jiang, Hadi Kharrazi, Rishi Saripalle, Vojtech Huser, Chunhua Weng, Nancy Roderer, Ana Szarfman, Niklas Elmqvist, David Gotz. Clinical Concept Value Sets and Interoperability in Health Data Analytics. AMIA Annual Symposium Proceedings. 2018 Dec 5;2018:480–489. PMCID: PMC6371254

7. Michelle R. Denburg, Hanieh Razzaghi, L. Charles Bailey, Danielle E. Soranno, Ari H. Pollack, Vikas R. Dharnidharka, Mark M. Mitsnefes, William E. Smoyer, Michael J. G. Somers, Joshua J. Zaritsky, Joseph T. Flynn, Donna J. Claes, Bradley P. Dixon, Maryjane Benton, Laura H. Mariani, Christopher B. Forrest, Susan L. Furth. Using Electronic Health Record Data to Rapidly Identify Children with Glomerular Disease for Clinical Research. Journal of the American Society of Nephrology: JASN. 2019 Dec;30(12):2427–2435. PMCID: PMC6900784

8. Michael G. Kahn, Tiffany J. Callahan, Juliana Barnard, Alan E. Bauck, Jeff Brown, Bruce N. Davidson, Hossein Estiri, Carsten Goerg, Erin Holve, Steven G. Johnson, Siaw-Teng Liaw, Marianne Hamilton-Lopez, Daniella Meeker, Toan C. Ong, Patrick Ryan, Ning Shang, Nicole G. Weiskopf, Chunhua Weng, Meredith N. Zozus, Lisa Schilling. A Harmonized Data Quality Assessment Terminology and Framework for the Secondary Use of Electronic Health Record Data. eGEMs. 2016 Sep 11;4(1):1244. PMCID: PMC5051581

9. Georgie May Massen, Philip W Stone, Harley H Y Kwok, Gisli Jenkins, Richard J Allen, Louise V Wain, Iain Stewart, Jennifer Kathleen Quint. Review of codelists used to define hypertension in electronic health records and development of a codelist for research. Open Heart. 2024 Apr 15;11(1):e002640. PMCID: PMC11029375

10. Joel Buchanan. Accelerating the Benefits of the Problem Oriented Medical Record. Applied Clinical Informatics. 2017 Feb 15;8(1):180–190. PMCID: PMC5373762

11. Joshua C. Feblowitz, Adam Wright, Hardeep Singh, Lipika Samal, Dean F. Sittig. Summarization of clinical information: a conceptual model. Journal of Biomedical Informatics. 2011 Aug;44(4):688–699. PMID: 21440086

12. Olivier Bodenreider, Duc Nguyen, Pishing Chiang, Philip Chuang, Maureen Madden, Rainer Winnenburg, Rob McClure, Steve Emrick, Ivor D’Souza. The NLM value set authority center. Studies in Health Technology and Informatics. 2013;192:1224. PMCID: PMC4300102

13. Adam Wright, Aileen P. Wright, Skye Aaron, Dean F. Sittig. Smashing the strict hierarchy: three cases of clinical decision support malfunctions involving carvedilol. Journal of the American Medical Informatics Association: JAMIA. 2018 Nov 1;25(11):1552–1555. PMCID: PMC6213087

14. Adam Wright, Joan S. Ash, Skye Aaron, Angela Ai, Thu-Trang T. Hickman, Jane F. Wiesen, William Galanter, Allison B. McCoy, Richard Schreiber, Christopher A. Longhurst, Dean F. Sittig. Best practices for preventing malfunctions in rule-based clinical decision support alerts and reminders: Results of a Delphi study. International Journal of Medical Informatics. 2018 Oct;118:78–85. PMCID: PMC6128667

15. Adam Wright, Thu-Trang T Hickman, Dustin McEvoy, Skye Aaron, Angela Ai, Jan Marie Andersen, Salman Hussain, Rachel Ramoni, Julie Fiskio, Dean F Sittig, David W Bates. Analysis of clinical decision support system malfunctions: a case series and survey. Journal of the American Medical Informatics Association. 2016 Nov 1;23(6):1068–1076.

16. Soumi Ray, Dustin S. McEvoy, Skye Aaron, Thu-Trang Hickman, Adam Wright. Using statistical anomaly detection models to find clinical decision support malfunctions. Journal of the American Medical Informatics Association: JAMIA. 2018 Jul 1;25(7):862–871. PMCID: PMC6016695

17. Adam Wright, Angela Ai, Joan Ash, Jane F Wiesen, Thu-Trang T Hickman, Skye Aaron, Dustin McEvoy, Shane Borkowsky, Pavithra I Dissanayake, Peter Embi, William Galanter, Jeremy Harper, Steve Z Kassakian, Rachel Ramoni, Richard Schreiber, Anwar Sirajuddin, David W Bates, Dean F Sittig. Clinical decision support alert malfunctions: analysis and empirically derived taxonomy. Journal of the American Medical Informatics Association. 2018 May 1;25(5):496–506.

18. Adam Wright, Trang T. Hickman, Dustin McEvoy, Skye Aaron, Angela Ai, Joan S. Ash, Jan Marie Andersen, Rachel Ramoni, Milos Hauskrecht, Peter Embi, Richard Schreiber, Dean F. Sittig, David W. Bates. Methods for Detecting Malfunctions in Clinical Decision Support Systems. Studies in Health Technology and Informatics. 2017;245:1385. PMID: 29295464

19. James. J. Cimino. Desiderata for Controlled Medical Vocabularies in the Twenty-First Century. Methods of information in medicine. 1998 Nov;37(4–5):394–403. PMCID: PMC3415631

20. James J. Cimino. In defense of the Desiderata. Journal of Biomedical Informatics. 2006 Jun;39(3):299–306. PMCID: PMC7185649

21. Alan L. Rector. What’s in a code? Towards a formal account of the relation of ontologies and coding systems. Studies in Health Technology and Informatics. 2007;129(Pt 1):730–734. PMID: 17911813

22. Rainer Winnenburg, Olivier Bodenreider. Issues in Creating and Maintaining Value Sets for Clinical Quality Measures. AMIA Annual Symposium Proceedings. 2012 Nov 3;2012:988–996. PMCID: PMC3540585

23. Rainer Winnenburg, Olivier Bodenreider. Metrics for assessing the quality of value sets in clinical quality measures. AMIA. Annual Symposium proceedings AMIA Symposium. 2013;2013:1497–1505. PMCID: PMC3900160

24. Licong Cui, Olivier Bodenreider, Jay Shi, Guo-Qiang Zhang. Auditing SNOMED CT hierarchical relations based on lexical features of concepts in non-lattice subgraphs. Journal of Biomedical Informatics. 2018 Feb;78:177–184. PMCID: PMC5835197

25. Licong Cui, Wei Zhu, Shiqiang Tao, James T. Case, Olivier Bodenreider, Guo-Qiang Zhang. Mining non-lattice subgraphs for detecting missing hierarchical relations and concepts in SNOMED CT. Journal of the American Medical Informatics Association: JAMIA. 2017 Jul 1;24(4):788–798. PMCID: PMC6080685

26. Fengbo Zheng, Jay Shi, Yuntao Yang, W. Jim Zheng, Licong Cui. A transformation-based method for auditing the IS-A hierarchy of biomedical terminologies in the Unified Medical Language System. Journal of the American Medical Informatics Association: JAMIA. 2020 Oct 1;27(10):1568–1575. PMCID: PMC7566369

27. O Bodenreider. Evaluating the Quality and Interoperability of Biomedical Terminologies. 2018 Apr.

28. A. L. Rector. Clinical terminology: why is it so hard? Methods of Information in Medicine. 1999 Dec;38(4–5):239–252. PMID: 10805008

29. M. Hassan Murad. Clinical Practice Guidelines: A Primer on Development and Dissemination. Mayo Clinic Proceedings. 2017 Mar;92(3):423–433. PMID: 28259229

30. Angelika Batta, Bhupinder Singh Kalra, Raj Khirasaria. Trends in FDA drug approvals over last 2 decades: An observational study. Journal of Family Medicine and Primary Care. 2020 Jan;9(1):105–114. PMCID: PMC7014862

31. Ronald J. Falk, Wolfgang L. Gross, Loïc Guillevin, Gary S. Hoffman, David R. W. Jayne, J. Charles Jennette, Cees G. M. Kallenberg, Raashid Luqmani, Alfred D. Mahr, Eric L. Matteson, Peter A. Merkel, Ulrich Specks, Richard A. Watts, American College of Rheumatology, American Society of Nephrology, European League Against Rheumatism. Granulomatosis with polyangiitis (Wegener’s): an alternative name for Wegener’s granulomatosis. Arthritis and Rheumatism. 2011 Apr;63(4):863–864. PMID: 21374588

32. M. A. Popovsky, M. D. Abel, S. B. Moore. Transfusion-related acute lung injury associated with passive transfer of antileukocyte antibodies. The American Review of Respiratory Disease. 1983 Jul;128(1):185–189. PMID: 6603182

33. Marc-Antoine Crocq. French perspectives on psychiatric classification. Dialogues in Clinical Neuroscience. 2015 Mar;17(1):51–57. PMCID: PMC4421900

34. Finale Doshi-Velez, Yaorong Ge, Isaac Kohane. Comorbidity clusters in autism spectrum disorders: an electronic health record time-series analysis. Pediatrics. 2014 Jan;133(1):e54–63. PMCID: PMC3876178

35. A. R. Feinstein. ICD, POR, and DRG. Unsolved scientific problems in the nosology of clinical medicine. Archives of Internal Medicine. 1988 Oct;148(10):2269–2274. PMID: 3140753

36. Anne Kveim Lie, Jeremy A. Greene. From Ariadne’s Thread to the Labyrinth Itself - Nosology and the Infrastructure of Modern Medicine. The New England Journal of Medicine. 2020 Mar 26;382(13):1273–1277. PMCID: PMC7919693

37. Jeffrey G. Klann, Lori C. Phillips, Alexander Turchin, Sarah Weiler, Kenneth D. Mandl, Shawn N. Murphy. A numerical similarity approach for using retired Current Procedural Terminology (CPT) codes for electronic phenotyping in the Scalable Collaborative Infrastructure for a Learning Health System (SCILHS). BMC medical informatics and decision making. 2015 Dec 11;15:104. PMCID: PMC4676189

38. Scott L. DuVall, Craig G. Parker, Amanda R. Shields, Patrick R. Alba, Julie A. Lynch, Michael E. Matheny, Aaron W. C. Kamauu. Toward Real-World Reproducibility: Verifying Value Sets for Clinical Research. Studies in Health Technology and Informatics. 2024 Jan 25;310:164–168. PMID: 38269786

39. Daniel Mendoza, Isca Amanda, Lin Zhao, Darwyn Chern, Maria Adela Grando. Ingredient-based method to create medication lists and support granular data segmentation. Health Informatics Journal. 2025;31(1):14604582251316781. PMID: 39873263

40. Sigfried Gold, Harold P. Lehmann, Lisa M. Schilling, Wayne G. Lutters. Value sets and the problem of redundancy in value set repositories. PloS One. 2024;19(12):e0312289. PMCID: PMC11627404

41. Nikolay Lukyanchikov, Kensaku Kawamoto. Evaluation of Discrepancies Among National Library of Medicine (NLM) Value Set Authority Center (VSAC) ICD-10-CM Value Sets: Case Study for Diagnoses of Common Chronic Conditions, Implications, and Potential Solutions. AMIA. Annual Symposium proceedings AMIA Symposium. 2023;2023:1087–1095. PMCID: PMC10785892

42. Bethany A. Van Dort, Wu Yi Zheng, Melissa T. Baysari. Prescriber perceptions of medication-related computerized decision support systems in hospitals: A synthesis of qualitative research. International Journal of Medical Informatics. 2019 Sep;129:285–295. PMID: 31445268

43. John D D’Amore, Laura K McCrary, Jody Denson, Chun Li, Christopher J Vitale, Priyaranjan Tokachichu, Dean F Sittig, Allison B McCoy, Adam Wright. Clinical data sharing improves quality measurement and patient safety. Journal of the American Medical Informatics Association. 2021 Jul 1;28(7):1534–1542.

44. Office of the National Coordinator for Health Information Technology. System Dashboard - ONC Jira [Internet]. [cited 2024 Jun 14]. Available from: https://oncprojectracking.healthit.gov/support/secure/Dashboard.jspa

45. Siru Liu, Aileen P. Wright, Barron L. Patterson, Jonathan P. Wanderer, Robert W. Turer, Scott D. Nelson, Allison B. McCoy, Dean F. Sittig, Adam Wright. Using AI-generated suggestions from ChatGPT to optimize clinical decision support. Journal of the American Medical Informatics Association: JAMIA. 2023 Jun 20;30(7):1237–1245. PMID: 37087108

46. James Shalaby. Can AI Replace Human Curation of Value Sets? [Internet]. Elimu Informatics. 2024 [cited 2024 Aug 6]. Available from: https://elimu.io/blog/can-ai-replace-human-curation-of-value-sets/

